# Preclinical and clinical validation of a novel injected molded swab for molecular assay detection of SARS-CoV-2 virus

**DOI:** 10.1101/2021.10.19.21265200

**Authors:** Chiara E. Ghezzi, Devon R. Hartigan, Justin Hardick, Rebecca Gore, Miryam Adelfio, Anyelo R. Diaz, Pamela D. McGuinness, Matthew L. Robinson, Bryan O. Buchholz, Yukari C. Manabe

## Abstract

During the COVID-19 public health emergency, many actions have been undertaken to help ensure that patients and health care providers had timely and continued access to high-quality medical devices to respond effectively. The development and validation of new testing supplies and equipment, including collection swab, help expand the availability and capability for various diagnostic, therapeutic, and protective medical devices in high demand during the COVID-19 emergency. Here, we report the validation of a new injection-molded anterior nasal swab, ClearTip™, that was experimentally validated in a laboratory setting as well as in independent clinical studies in comparison to gold standard flocked swabs. We have also developed an *in vitro* anterior nasal tissue model, that offers an efficient and clinically relevant validation tool to replicate with high fidelity the clinical swabbing workflow, while being accessible, safe, reproducible, time and cost effective. ClearTip™ displayed a greater efficiency of release of inactivated virus in the benchtop model, confirmed by greater ability to report positive samples in a clinical study in comparison to flocked swabs. We also quantified in multi-center pre-clinical and clinical studies the detection of biological materials, as proxy for viral material, that showed a statistically significant difference in one study and a slight reduction in performance in comparison to flocked swabs. Taken together these results underscore the compelling benefits of non-absorbent injected molded anterior nasal swab for COVID-19 detection, comparable to standard flocked swabs. Injection-molded swabs, as ClearTip™, could have the potential to support future swab shortage, due to its manufacturing advantages, while offering benefits in comparison to highly absorbent swabs in terms comfort, limited volume collection, and potential multiple usage.

## Introduction

The emergence of SARS-CoV-2 and the subsequent COVID-19 pandemic has resulted in approximately 200 million cases globally (1). The long incubation period as well as the high prevalence of asymptomatic carrier transmission are prominent factors for the high transmission rate of SARS-CoV-2 (2). Thus, efficient and reliable detection of infectious carriers through large-scale surveillance is critical to limit further spreading of the disease, particularly as many countries experience multiple waves of infections. Despite effective vaccines, continued global propagation of variants with efficient transmission dynamics even in vaccinated hosts increases the need to provide easily accessible testing on a wide variety of platforms.

A nasopharyngeal (NP) swab is the gold standard sampling method to detect SARS-CoV-2, as recommended by the World Health Organization (WHO) (3). Sample collection with NP swab is performed by trained professionals, transferred into transport media, and then processed via RNA purification and RT-qPCR, or other FDA approved methods. NP swabs are 15 cm long to reach the posterior nasopharynx, with a head diameter between 1 to 3.2 mm made from short synthetic threads, flock, or spun fibers (4). The NP collection process is often reported as uncomfortable (5). Due to supply chain limitations and increased demand, anterior nasal (AN) specimens have been successfully adopted as an effective alternative to solve the NP swab shortage (6), and nasal swabs have been found to be equivalent to NP swabs for the detection of SARS-CoV-2, as studies have indicated high sensitivity and specificity for that sample type (7). AN swabs offers similar testing sensitivity to NP swabs, while they are easier to administer and more comfortable for the patients, supporting its use in large-scale screening testing (5).

New swab production methods have been explored to address the increase in demand for nasal swabs. 3D printed and injected molded swabs displayed performance comparable to standard flocked swabs (4, 8), while offering high throughput, cost effective alternatives (8, 9). In addition, a non-absorbent swab would allow to collected samples in limited volumes and to elute them into smaller volume of transport media, in comparison to standard absorbent swab, resulting in greater sample concentration and subsequent increase in sensitivity in viral RNA detection.

To validate new swabs, innovative experimental procedures are needed to streamline this process. We evaluated the performance of the novel ClearTip™ (Yukon Medical, Durham, NC) injected-molded swab in comparison to traditional flocked nasal swabs. Preclinical validation was performed with an innovative *in vitro* tissue model to mimic the soft tissue structure as well as the viscous mucous component of the nasal passage. We hypothesized that the simulation of such tissue properties will provide a bench top validation to further support subsequent clinical studies. Finally, we evaluated the performance of the ClearTip™ injected-molded swab in comparison to traditional flocked nasal swabs in accurately collecting patient samples as well as detecting SARS-CoV-2 in two independent clinical studies.

## Materials and Methods

### Materials

Anterior nasal, mid-turbinate, and nasopharyngeal injected molded swabs were designed, produced, and manufactured by Yukon Medical (Figure 1). These swabs were then compared to commercially available flocked swabs (Steripack and Copan).

**Figure 1.**
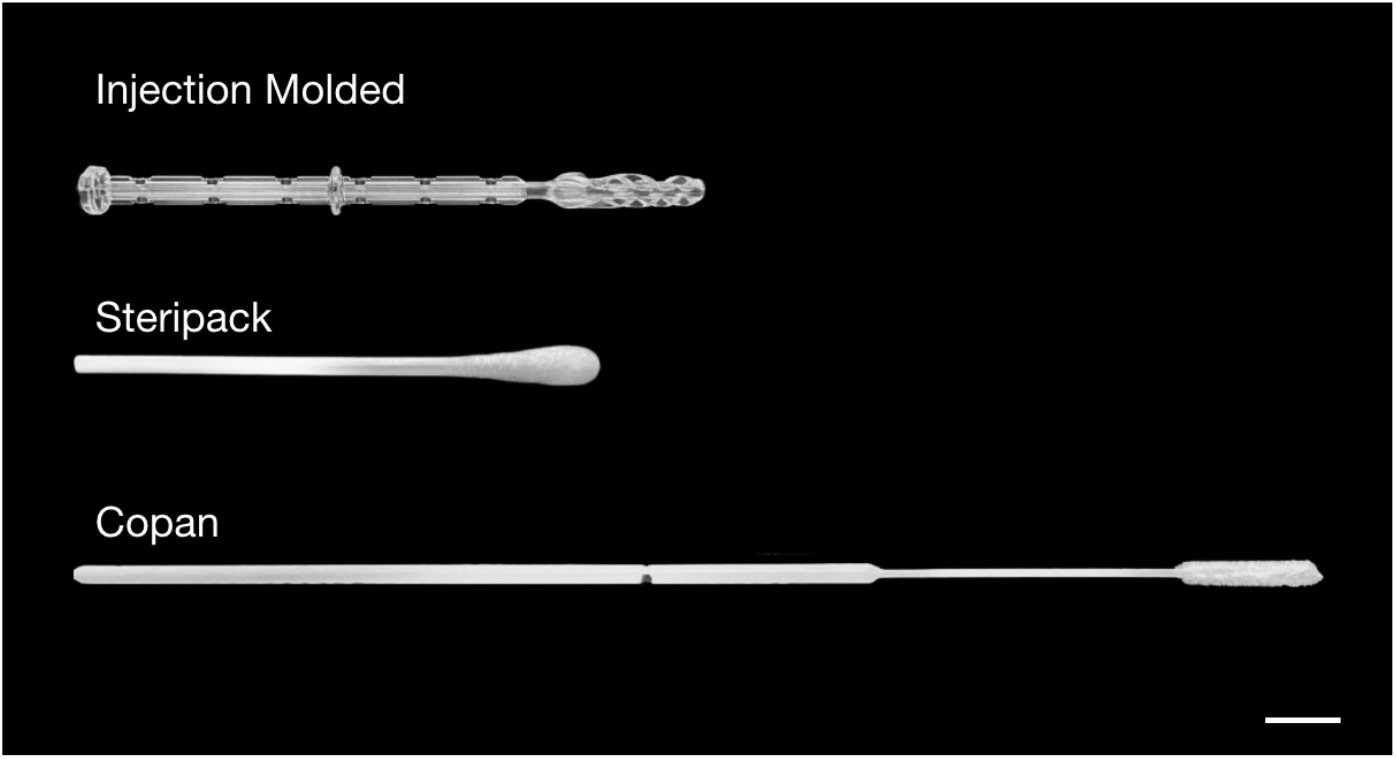
Experimental swabs. Macro images of Injection-molded ClearTip™, Steripack, and Copan anterior nasal swabs used in these studies. Scale bar = 10 mm.

### Preclinical Validation

#### 1. Anterior Nasal Tissue Model Preparation

A nasal tissue model was developed to perform *in vitro* pre-clinical studies to quantify swab pick-up and release of inactivated SARS-CoV-2 virus from a physiologically relevant nasal mucosa. The nasal tissue model was comprised of a natural sponge made of cellulose (7456T-C41, 3M, Saint Paul, MN, USA) to mimic the soft tissue nature. It was prepared with hollow punchers in a cylindrical structure with an external and internal diameter of 2.5 and 0.875 mm, respectively, and 26 mm in overall length. To confine and retain the mucus, the tissue model was inserted in a polyvinyl chloride external tubing (Everbilt). The sponge was disinfected in subsequent washes of 10% bleach, 70% ethanol, deionized water, and then autoclaved. Before mucus saturation, the model was left to dry overnight is a biological hood. The mucus mimicking solution was prepared from a 2 wt% polyethylene oxide solution (CAS#: 25322-68-3 Acros Organics LOT#: A0403177), that has been previously shown to have similar viscosities to healthy nasal mucus (10, 11). The anterior nasal tissue model was then saturated with 4.5 ml of the physiologically relevant mucus solution.

#### 2. Pick Up Swab Quantification

To quantify swab uptake, the anterior nasal tissue model was saturated with the physiologically relevant mucus solution and the following swabbing procedure was utilized. Each swab was inserted into the model until resistance was encountered, twisted around the nasal model surfaces five times, held in place for 15 seconds, and then removed. The pick-up swab quantification was performed by gravimetrical analysis for the ClearTip™ NP and ClearTip™ MT/N swabs in comparison to commercially available flocked swabs (Steripack), and the weight of each swab (N=5) was recorded before and after the swabbing procedure, and reported as mass uptake, for three independent experiments.

#### 3. Swab Release Quantification

To quantify swab release, the anterior nasal tissue model was saturated with the physiologically relevant mucus solution spiked at the concentration of 10^6^ copies/ml of heat-inactivated SARS-CoV-2, USA-WA1/2020 (NR-52286, BEI Resources, ATCC, USA) and the swabbing procedure in the bench model was performed, as described above. After the procedure, the swab was removed and placed into an Eppendorf tube with 350 μl of viral transport media (VTM). The vial with the swab was then vortexed for 30 sec, sonicated for 1 min, and vortexed again for 30 sec. 5 μl from each sample was then tested to quantify the detection of SARS-CoV-2, as an expression of pick up and release of the different swabs. To evaluate the presence of SARS-CoV-2, we performed the CDC 2019-Novel Coronavirus (2019-nCoV) Real-Time RT-PCR Diagnostic Panel (https://www.fda.gov/media/134922/download), per manufacturer instructions using the 2019-nCoV_N1 Combined Primer/Probe Mix with a Quantabio qScript XLT One-Step RT-qPCR ToughMix. Amplification was performed following manufacturer instructions with a QuantStudio™ 5 Real-Time PCR System (Thermo Fisher Scientific, Waltham, MA, USA). The results for each swab (N=5) were reported as cycle threshold (Ct) value.

#### 4. Preclinical Human Sampling

Laboratory staff were approached regarding volunteering for a self-sampling study. Volunteers (N=8) were provided with one Copan FLOQSwabs 220250 Regular, one ClearTip™ swab and two 15ml conical tubes containing 3ml of VTM. Each volunteer was asked to utilize the flocked swab in their right nasal cavity and the ClearTip™ in their left nasal cavity. Sampling for both swab types was performed identically by inserting the swab until resistance was noticed, the swabs were rotated 5 times, and then removed and placed into the separate tubes of VTM. Following swabbing, each tube of VTM was utilized to perform cell counts with the Countess II (Life Technologies, Foster City, CA). Briefly, 10 μl of VTM from each tube was mixed with 10 μl Trypan Blue (Life Technologies), and 10 μl of the mixture was loaded into a Countess II slide. Cell counts were performed utilizing the Countess II default setting. Following cell counting, 200 μl of VTM from each tube was extracted for RT-qPCR analysis utilizing the Nuclisense EasyMag (Biomeriuex, Durham, NC) following manufacturer instructions. Post-extraction, each sample was analyzed for total DNA concentration utilizing the NanoDrop One (Thermofisher Scientific, Waltham, MA) per manufacturer instructions. Samples were subsequently analyzed for RNAseP utilizing the CDC nCOV_2019 RT-qPCR assay per manufacturer instructions. Cell counts, total DNA concentration and RNASeP Ct values were utilized to discern differences in the quantity of cells released by each swab type.

### Clinical Validation

To evaluate ClearTip™ versus flocked swab performance, two clinical studies were carried out to assess pick-up and release of each swab type by quantifying human RNase P expression, as well as N1 and N2 when the subjects were positive.

#### 1. Clinical Study I

##### 1.1. Study design and Oversight

Participants were already enrolled into a surveillance program at University of Massachusetts Lowell (UML), and were asked to be swabbed with the ClearTip™ N swab as well as with a commercially available comparator (Steripack), and to complete a short survey on the comfort of the swabs. Swabs were collected in VTM in a 15-ml conical tube, transported to the RADx Validation Core Lab at UML, stored at 4°C identically to clinical samples, and tested the following day. Sample collection was performed by trained nursing personnel at the UML Wellness Center. This study was reviewed and approved by the institutional review board of UML (protocol number 20-149-BUC-FUL).

##### 1.2. Participants

Participants were healthy individuals, as self-reported and determined by a screening asked during recruitment consent, and already enrolled into the surveillance program at UML. Patient AN samples (N=47) were collected with a ClearTip™ swab within seven days of a validated negative SARS-CoV-2 test result from a sample collected with a CLIA use approved Class I exempt nasal swab and tested in an approved RT-qPCR assay by a CLIA-certified clinical lab. Adults over 18 years of age were given a participant information sheet by study staff and asked whether they would agree to being swabbed with an experimental swab by a trained nurse to the control swab required for testing, and also complete a short comfort survey.

##### 1.3. Study procedures

ClearTip™Injected molded swabs were individually packaged and autoclaved for sterilization according to manufacturer protocols. Swabbing was performed per the standard protocol in both nostril for each swab. After swabbing both nostrils, the subject returned to the study coordinator(s). Participants then completed a short survey on the relative comfort of the two swabs. They rated each swab on 5-point Likert scale with 1=very uncomfortable and 5=very comfortable. The order of swabbing with the control or the experimental swabs was random for all the participants. Control and experimental swabs were placed in separate vials of VTM and transported to the RADx Validation Core Lab, where each sample was tested on the QuantStudio™ 5 Real-Time PCR System with 2019-nCoV_N1, 2019-nCoV_N2 and Human RNase P Combined Primer/Probe Mix with a Quantabio qScript XLT One-Step RT-qPCR ToughMix per the standard CDC protocol.

#### 2. Clinical Study II

##### 2.1. Study design and Oversight

Participants, enrolled in the study performed at Washington University School of Medicine, were recruited as symptomatic at a walk-in clinic. They were all tested first with NP swabs and then randomly swabbed nasally first with the ClearTip™ N swab or commercially available comparator (Copan). Swabs were eluted for 10 minutes in M4-RT universal transporting medium (UTM). Sample collection was performed by trained nursing personnel at the Washington University School of Medicine. This study was reviewed and approved by the institutional review board of Washington University School of Medicine (protocol number 7453).

##### 2.2. Participants

Participants were symptomatic individuals, as self-reported and determined by a screening asked during recruitment consent, with a confirmed EUA approved RT-PCR COVID-19 virus test result, performed with nasopharyngeal (NP) flocked swabs processed with Cepheid Xpert® Xpress SARS-CoC-2. Patient AN samples (N=38) were collected with a ClearTip™ swab in comparison to flocked swabs (FLOQSwabs™ Contoured Adult, Copan). Adults over 18 years of age were given an informed consent form by study staff and asked whether they would agree to being swabbed with an experimental swab by a trained nurse to the control swab required for testing.

##### 2.3. Study procedures

ClearTip™ swabs were individually packaged and autoclaved for sterilization according to manufacturer protocols. Two paired anterior nares nasal swabs from each nare were collected from each enrolled subject. After swabbing both nostrils, the subject returned to the study coordinator(s). The order of swabbing with the control or the experimental swabs was random for all the participants. Control and experimental swabs were placed into the M4-RT UTM and shipped to shipped to the Microbiology Lab at the Grady Memorial Hospital (Atlanta, GA, USA), stored at 4°C identically to clinical samples, and tested the following day. Samples were tested on the QuantStudio™ 5 Real-Time PCR System with 2019-nCoV_N1, 2019-nCoV_N2 and Human RNase P Combined Primer/Probe Mix with a Quantabio qScript XLT One-Step RT-qPCR ToughMix per the standard CDC protocol.

### Statistical Analyses

Paired t-tests and equivalence testing using a two one-sided t-test (TOST) of equivalence using appropriate equivalence margins were performed with a p<0.05 significance threshold (12). Negative predictive value (NPV) was calculated with a weighted generalized score method (13). Paired t-test, equivalence tests and the weighted generalized score method were analyzed using SAS 9.4 (SAS Institute, Inc., Cary NC). Additional Student’s T-test (T-test) with a p-value of < 0.05 was performed with Origin(Pro), Version 2021b OriginLab Corporation, Northampton, MA, USA.

## Results

### Benchtop validation

An anterior nasal tissue model was developed to quantify the sample uptake and release from anterior nasal swabs (Figure 2A). The tissue model was designed with a cellulose sponge to mimic the nasal soft tissue, and saturated with a synthetic nasal mucus spiked with inactivated SARS-CoV-2 virus and was designed to mimic clinical nasal swab to facilitate an evaluation of swab performance. The swab uptake was quantified by gravimetric analysis after the benchtop swabbing procedure. The ClearTip™ swab displayed a significantly lower uptake of synthetic mucus in comparison to control flocked swabs (p<0.05). ClearTip™ demonstrated more than 20-fold reduction in swab uptake in comparison to the control swab (Figure 2B). The ability of the swabs to release biological materials was quantified by performing the swabbing workflow with the nasal tissue model loaded with inactivated SARS-CoV-2 virus. Swabs were then collected into viral transport media, and the virus released from the swab was quantified via RT-qPCR, according to CDC guidelines. ClearTip™ displayed a significantly lower cycle time (e.g., more virus) in detecting inactivated SARS-CoV-2 virus, in comparison to flocked control swab (p<0.05). The reduction in cycle time for the ClearTip™ swab support greater viral recovery in comparison to flocked control (Figure 2C).

**Figure 2.**
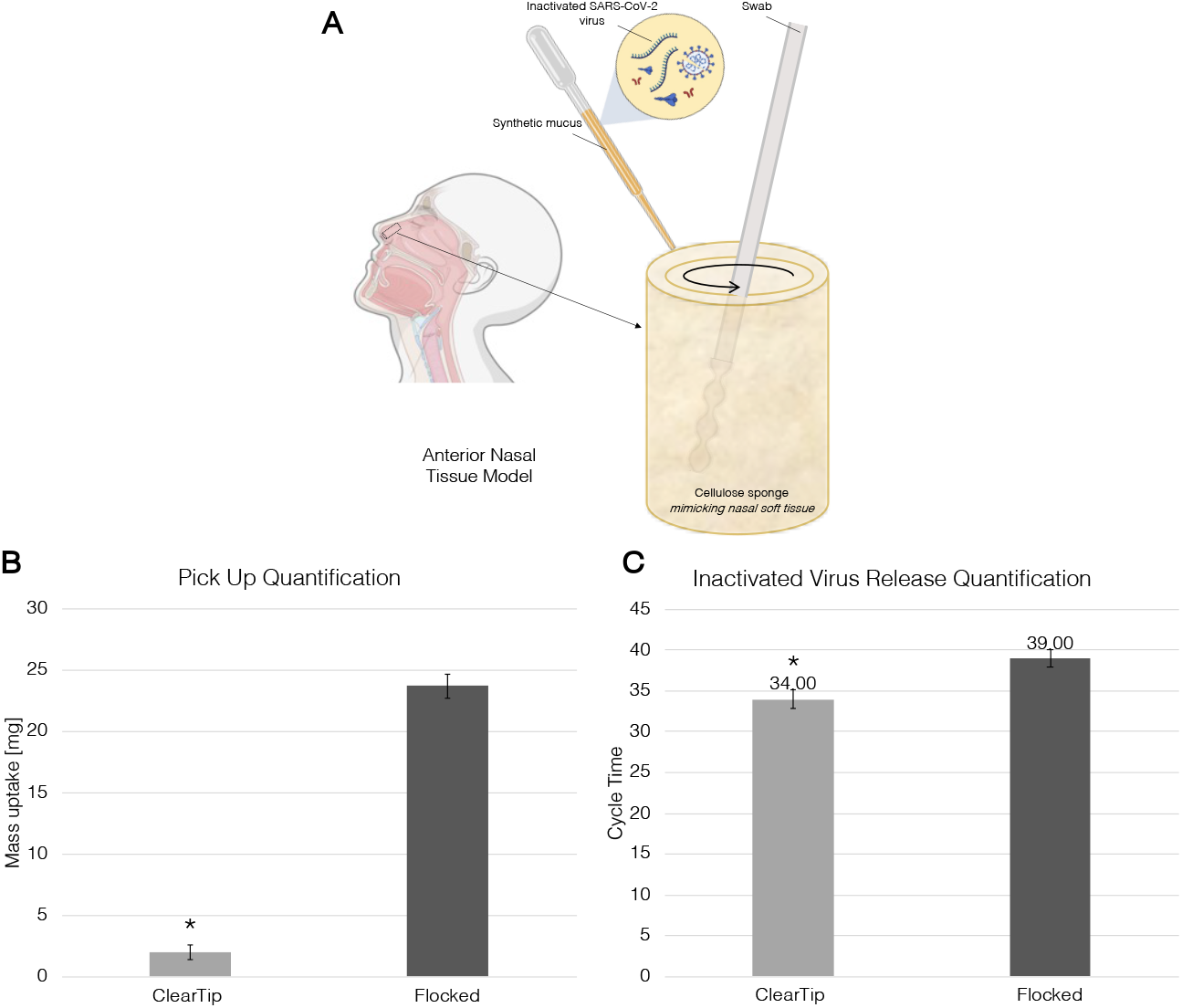
ClearTip™ benchtop validation. A. Anterior nasal tissue model comprised of a cellulose sponge loaded with synthetic nasal mucus spiked with inactivated SARS-CoV-2 virus. The cellulose sponge would mimic the nasal soft tissue during the bench top swabbing procedure with the ClearTip™ in comparison to flocked control swab. B. Mass uptake during the benchtop swabbing procedure was quantified by gravimetrical analysis. Flocked control samples displayed statistically greater mass update compared to ClearTip™. C. Swab material release was collected into viral transport media and inactivated virus was quantified using PCR following CDC guidelines. * Statistical difference against flocked control group (p<0.05). Created with BioRender.com.

### Preclinical Human Sampling

Results from the preclinical self-sampling study are summarized in Figure 3. Numbers of total cells, live cells and dead cells were quantified, as well as the quantification of DNA content and RNase P expression. Overall, samples that were from self-sampling with flocked swabs produced higher live cell counts (Figure 3B), greater DNA concentration (Figure 3D) and lower RNAseP Ct values (Figure 3E) than the samples from ClearTip™ self-sampling (p<0.05). Quantification of the total number of cells (Figure 3 A) and dead cells (Figure 3C) were not statistically significantly different at the 0.05 level. While the t-test found non-significant differences (p=0.0509 and p=0.2920, respectively for total and dead cells), the underpowered sample size and large variation contributed to the non-equivalence result, from the equivalence test.

**Figure 3.**
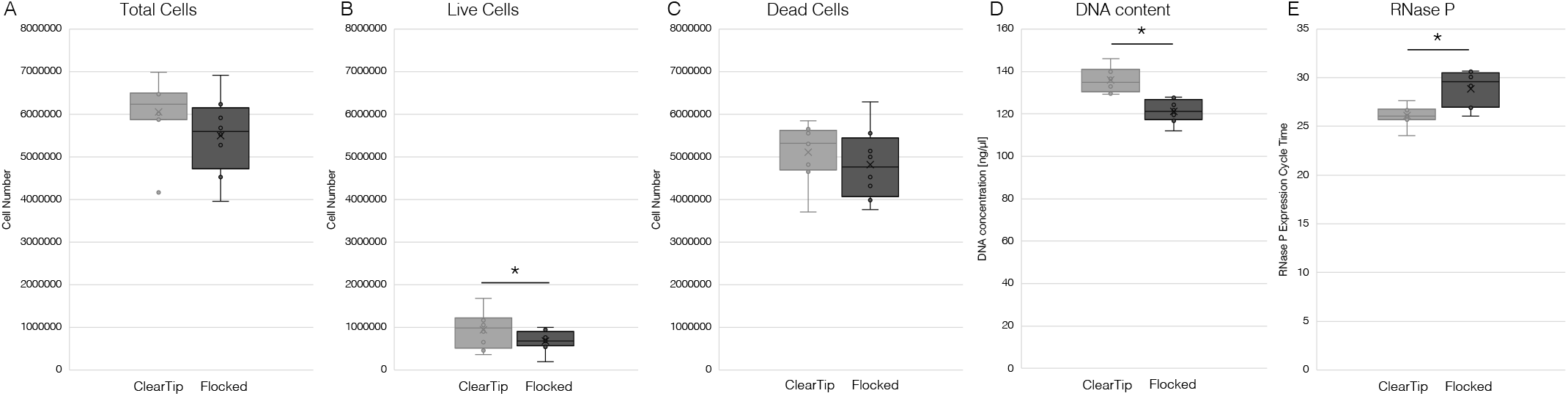
ClearTip™ Preclinical self-sampling validation. Pre-clinical self-sampling study was performed to compare performance of ClearTip™ swab against flocked swab. In this study, numbers of total cells (A), live cells (B), and dead cells (C) were quantified, as well as DNA content (D) and RNase P expression (E). Statistically significant differences were reported only for live cells, DNA content and RNase P (p<0.05).

### Clinical validation

To quantitively compare the performance of the ClearTip™, 47 participants self-collected nasal samples using ClearTip™ in comparison to a flocked CLIA approved swab (Figure 4A). None of the participants tested positive in this surveillance program, thus, the expression of RNase P was used to compare the efficiency of each swab to pick up human biological material. ClearTip™ demonstrated a significantly higher RT-qPCR cycle time in comparison to the flocked control swab, Steripack (p<0.05). Quantitatively, we reported a 2-cycle difference for the two compared swabs, that was found statistically different.

**Figure 4.**
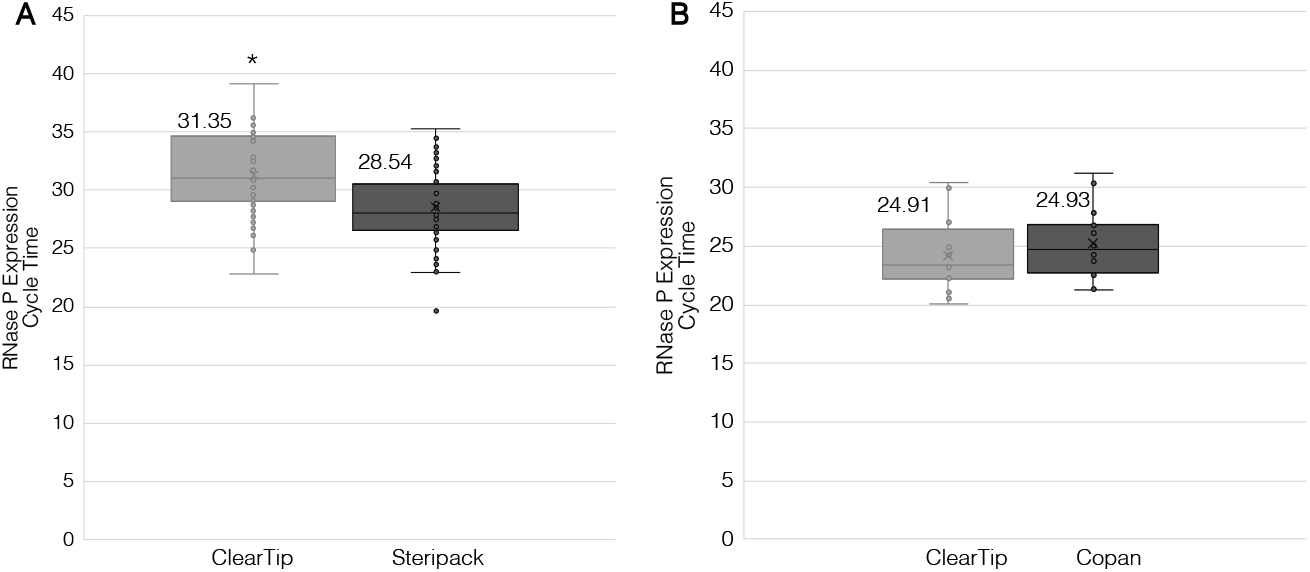
ClearTip™ clinical validation for RNase P detection. Two independent clinical studies were performed to quantify the ability of the ClearTip™ swab to pick up nasal cellular material in comparison to control flocked nasal swabs. A. Clinical study at CAPCaT University of Massachusetts Lowell showed a significant increase in cycle time for detection of RNase P collected by ClearTip™ Swabs and Steripack Nasal Swabs (p<0.05). B. Clinical study at Washington University St. Louis showed no statistical difference (p=0.945) in cycle time for detection of RNase P collected by ClearTip™ Swabs and Copan Nasal Swabs. * Statistical difference against flocked NP control (p<0.05).

In the second clinical study, 38 participants were collected using ClearTip™ in comparison to a flocked CLIA approved swab (Figure 4B). ClearTip™ displayed no differences in comparison to the CLIA approved swab, Copan (p=0.9549). In addition, an equivalence test with a margin of 1 determined that the swabs measured the same value of RNAseP. Using an RNAaseP RT-PCR, there was no difference between ClearTip™ and the Copan swabs. Of the 9 individuals (of 38 total) who tested NP swab positive in a CLIA-certified reference lab (Figure 5), the ClearTip™ swab confirmed 78% of positive cases detected with an NP CLIA use approved flocked swab for N gene detection, while the Copan swab confirmed only 22% of the cases. Based on a weighted generalized score method, ClearTip™ displayed an NPV of 90%, while the comparator swabs showed and NPV of 79% (p<0.05).

**Figure 5.**
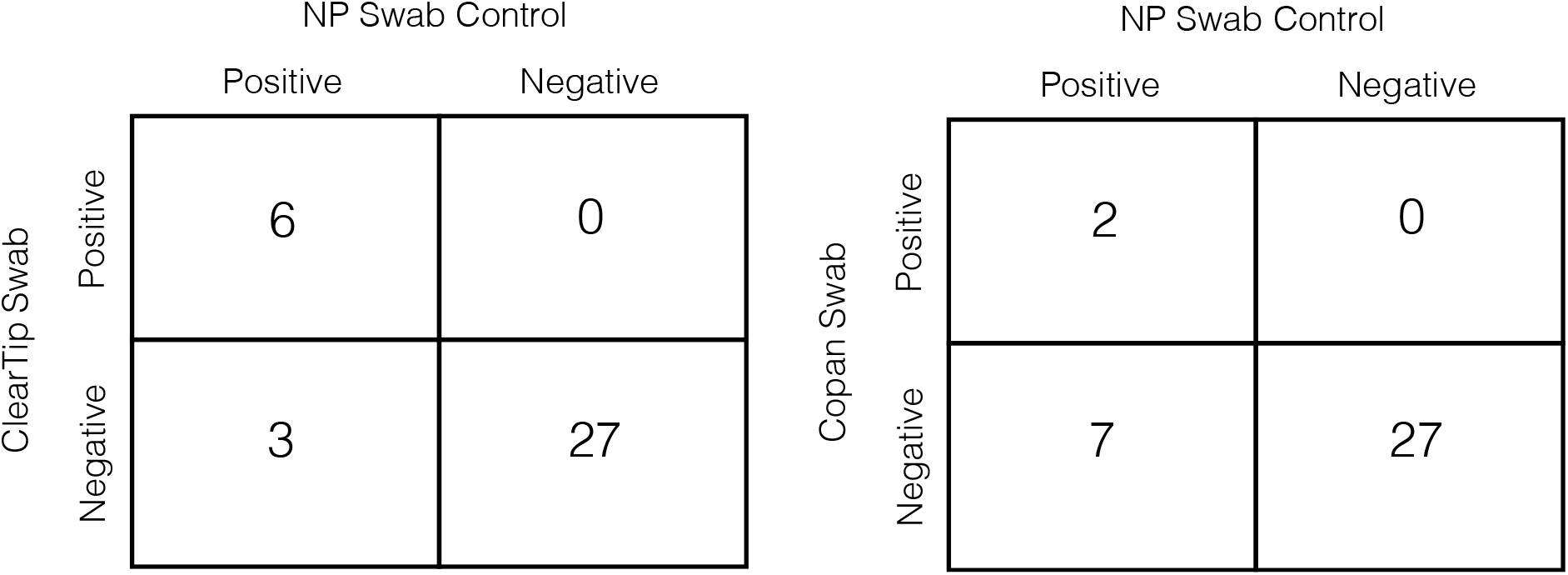
ClearTip™ concordance with control swabs. Summary of positive and negative samples detected with ClearTip™ swab in comparison to Copan flocked swab. Status of participants enrolled in the study were confirmed by EUA approved RT-PCR COVID-19 virus test result, performed with NP flocked swabs.

In the first clinical study, participants scored the comfort of the two swabs over a 5-point Likert scale (Figure 6A). The mean paired difference in swab comfort was reported 0.3 (Figure 6B). ClearTip™ was found equivalent to the flocked CLIA approved swab with a paired two one-sided t-test (TOST) of equivalence using an equivalence margin of 1 (p<0.05).

**Figure 6.**
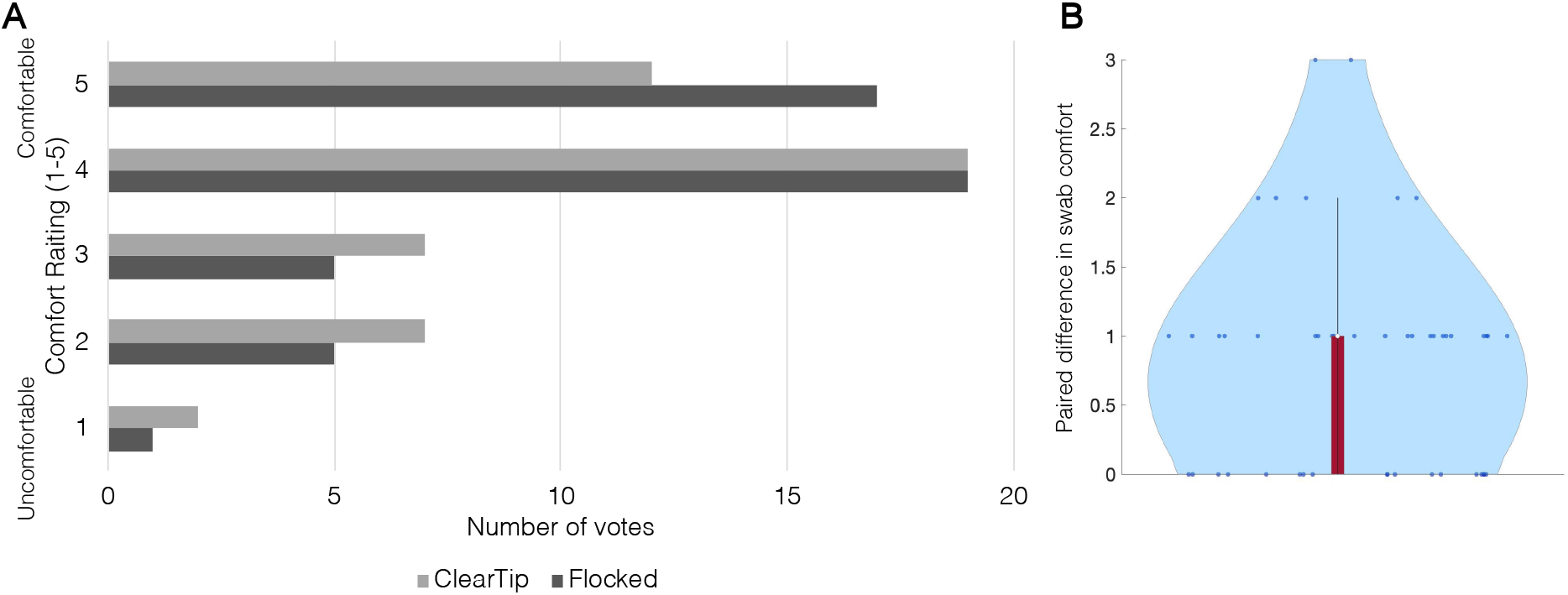
ClearTip™ comfort study. A. Summary of comfort survey with ratings on 5-point Likert scale with 1=very uncomfortable and 5=very comfortable. B. Mean paired difference in swab comfort. ClearTip™ was found equivalent to the flocked CLIA approved swab with a paired t-test equivalence margin of 1 (p<0.05).

## Discussion

During the peaks in the pandemic, swab shortages at hospital across the United States and worldwide have been reported (14), prompting the need to develop new swab prototypes with comparable performance and capable of high-volume manufacturing, while being cost-efficient. Here, we report a new injection-molded anterior nasal swab, ClearTip™, that was experimentally validated in a laboratory setting as well as in independent clinical studies in comparison to gold standard flocked swabs.

In vitro tissue models have been recently developed and extensively investigated, as they can mimic a wide range of the physical, structural, and biological characteristics of native tissues, and thus implemented to study *in vitro* physiological and pathological tissue conditions (15-19).

We report an anterior nasal tissue model, that mimics the architecture and structure of the native soft tissue, as well as the physical properties of the nasal mucus. In the anterior nasal tissue model, we quantified the ability of ClearTip™ to pick up artificial mucus in comparison to gold standard flocked swabs. ClearTip™ displayed more than 20 times less retention than a flocked swab. However, when we evaluated *in vitro* the capacity of ClearTip™ to release viral material, the injected molded swab displayed significantly greater inactivated virus release in comparison to flocked swabs. This suggests a much greater efficiency of release of inactivated virus from the ClearTip™ swab, that may offset the reduction in uptake in comparison to standard flocked swab.

In clinical assessments, ClearTip™ showed variability in its ability to pick up and release cellular material as a proxy for performance in COVID-19 testing. Measurements of cell population, DNA concentration and RNaseP quantification showed equivalence in one study and a reduction in performance in comparison to flocked swabs in a separate assessment suggesting that the protocol for use may make a difference in the ability of the injection-molded swab to pick up cells and, therefore, virus. In a small clinical assessment, flocked and ClearTip™ nasal swabbing detected 22% and 78% of NP swab SARS-CoV-2 positive paired samples.

The *in vitro* anterior nasal tissue model offers an efficient and clinically relevant validation tool that allows to replicate with high fidelity the clinical swabbing workflow, while being accessible to researchers, safe, reproducible, time and cost effective. We also demonstrated the concordance of the *in vitro* model results with our clinical studies. The non-absorbent injection-molded ClearTip™ swab could have the potential to support future swab shortage, due to its manufacturing advantages, while offering benefits in comparison to highly absorbent swabs in terms comfort, limited volume collection, and potential multiple usage.

Injection-molded swabs can be mass produced at limited costs from well-characterized polymeric materials, while leveraging years of experience in medical devices manufacturing from injection molding processes. In addition, the non-absorbent head and superior material release may allow greater sensitivity when used in pooling studies, though the less efficient capture of cells with the current instructions for use may require optimization of technique. The single solid material design could potentially allow multiple use of the same injected molded swab, after cleaning, disinfection and sterilization, under extenuating circumstances, as several low- and middle-income countries faced in the midst of the COVID-19 pandemic.

## Data Availability

All data produced in the present study are available upon reasonable request to the authors

## Acknowledgements

This study was funded by the NIH RADx-Tech program under 3U54HL143541-02S1, 3U54HL143541-02S2 and U54EB007958-12S1. The views expressed in this manuscript are those of the authors and do not necessarily represent the views of the National Institute of Biomedical Imaging and Bioengineering; the National Heart, Lung, and Blood Institute; the National Institutes of Health, or the U.S. Department of Health and Human Services. Salary support to YCM and JH from U54EB007958 (Center POC Technologies Research for Sexually Transmitted Diseases).

